# Influencers of effective behavior change communication interventions delivered by community health workers in adults: A Scoping Review Protocol

**DOI:** 10.1101/2020.11.29.20240432

**Authors:** Raunaq Singh Nagi, Pankaj Prasad, Sanjeev Kumar

**Affiliations:** Department of Community and Family Medicine, All India Institute of Medical Sciences, Bhopal

**Keywords:** BCI, lay health workers, adults, modifiable risk factors, LMICs

## Abstract

**Introduction:** Behavior Change Communication (BCC) serves as a key pathway for delivery of messages for modifying risky behaviors such as unsafe sex, tobacco use, consumption of unhealthy diet and sedentary lifestyle. Behavior Change Communication has been successfully applied in various health conditions, settings and on different participants. In Low- and Middle-Income Countries (LMICs), the delivery of BCC is achieved through Community Health Workers (CHWs) due to limited availability of medical personnel. Current evidence indicates that delivering such interventions through CHWs is a promising approach to achieve desired behavior change and has potential to be upscaled. However, unavailability of information regarding the applicability of these interventions at different community settings, health conditions, and medium for intervention delivery, has made upscale and implementation a challenge. This scoping review will summarize the scope of settings, communication channels, and characteristics of message delivery protocols of behavior change communication interventions targeted at adults delivered via CHWs.

**Methods and analysis:** The scoping review methodology framework outlined by Arskey and O’Malley will guide this review. We will search the following databases, MEDLINE, ERIC, JSTOR, ScienceDirect, using pre-defined search strategy. We will include studies published in English language, without any limits on the time of publication. Firstly, titles and abstracts will be screened, followed by full-length articles, for inclusion in the review. We will extract the data in a well-defined template developed for the purpose. All the reviewers will synthesize the evidence regarding and present the results using descriptive statistics and narrative.

**Ethics and dissemination:** This review is being conducted as a part of a doctoral thesis approved by the institutional ethics committee. The results of this scoping review will be disseminated in the form of peer-reviewed publication, and presented in conferences and will be used to design behavior change intervention to be introduced in community.

**Strengths and limitations:**

- This will be the first scoping review to scale the community settings where behavior change communication interventions have been delivered.
- This review will also scale the characteristics of such interventions, viz, modality and medium of communication, and duration and periodicity of interventions.
- This review will only include articles published in English language across the named freely searchable databases.
- Assessment of quality of the included studies is beyond the scope of this review and hence will not be carried out.

## Introduction

Health behavior, as described by Cockerham (1) is the activity individuals undertake to maintain or enhance their health, prevent health problems, or achieve a positive body image. Contrastingly, certain behaviors, known as behavioral risk factors, have been linked to compromised health. Insufficient nutritional intake among children and mothers, tobacco use, alcohol and other substance abuse, excess intake of certain unhealthy dietary constituents, sexual abuse and violence, unsafe sex and inadequate physical activity are a few such behavioral risk factors. (2) These behavioral risk factors lead to altered physiological states and metabolic risk factors. Combined with genetic risk factors of an individual, they lead to development of several non-communicable diseases (NCDs). (3–6)

Several theories have been postulated to understand health behaviors, and describe the process of developing a particular health behavior. These theories are also used to devise strategies to modulate these risky health behaviors (7,8) Health Belief Model (HBM) (9) has been used by researchers to design facility-based osteoporosis prevention programs in Iran (10), and community-based educational intervention for adoption of skin cancer-prevention behavior among farmers. (11) Transtheoretical Model (TTM) (12) has been used to understand the smoking behavior of adolescents in educational setting (13), explain physical activity self-efficacy among African-American adolescents (14), and even to devise interventions to manage depression in hospitalized patients of coronary artery disease in China. (15) And, Theory of Reasoned Action (TRA) (16) has been used to identify perceptions on teenage pregnancies among American Indians (17), devise school-based interventions to promote breakfast consumption among Iranian children (18) and design self-care facility-based intervention for diabetic Iranian women. (19) Similarly, Theory of Planned Behavior (TPB) and Social Cognitive Theory (SCT) have also been applied to various domains of behavior change such as physical activity promotion. (20,21) Furthermore, more than of these theories have been combined and used by researchers, such as for identification of individuals seeking healthcare and subsequently taking appropriate actions for them. (22) Recent studies have demonstrated success of community- or peer-guided support planned using theory-based interventions in addition to personalized interventions. (23,24)

These theory-based approaches for behavior change are termed ‘behavior change interventions’ (BCI). When applied in community-based settings, BCIs require public health communication. Public health communication, also known as behavior change communication (BCC) is defined as a transdisciplinary, audience-centred, change-oriented, multi-level communication strategy. It targets a variety of audience and conglomerations. (25) BCIs and public health communications have been designed and delivered as mass media campaigns (26), media advocacies (27), social marketing (28), targeted messages (29) and tailored messages. (30)

Applications of BCC in inducing sustained behavior change have been identified globally across a variety of health conditions, such as maternal-child health, nutrition, vector-borne diseases, sexually transmitted diseases and others. (31–34). These interventions have demonstrated applicability across various age groups and ease with the use of different modes of communication for delivery, such as telephonic surveys in improving women’s pre-conceptional health in Pennsylvania (35), telephonic intervention by physical therapists in post-surgical spinal rehabilitation (36), or internet-delivered personalized nutrition intervention across seven countries in Europe. (37) Hence, these communication interventions are universally applicable and effective.

In Low- and Middle-Income Countries (LMICs), where resources and availability of trained medical personnel are limiting factors for healthcare delivery, community health workers (CHWs; identified by other names such as lay health workers, volunteer health workers and others) liaison public and healthcare, and play a vital role in healthcare delivery. In such countries, the onus of delivering successful BCC also lies with the CHWs. And the potential of CHWs in delivery of such interventions has already been recognized. (38) Studies have demonstrated that trust in CHWs also plays a crucial role in the success of an intervention, further highlighting their significance in community-based BCCs. (39,40)

As the body of evidence pertaining to the role of CHWs in BCC delivery continues to grow, we have proposed to undertake a scoping review to identify various characteristics of behavior change communication strategies, namely, community-based settings they have been delivered in, channels of communication that have been used, and message delivery protocols, that might influence the success of the intervention.

## Methods and analysis

### Protocol design

The review is structured on the methodological background laid down by Arksey and O’Malley (41) and the recommendations JBI methodology of scoping reviews. (42)

For reporting the results, this review will use the Preferred Reporting Items for Systematic reviews and Meta-Analyses extension for Scoping Reviews (PRISMA-ScR) guidelines and checklist. (43)

The individual steps of the scoping methodology are elaborated below.

#### 1. Identification of research question

The Population/Participant (P), Concept (C), and Context (C) (PCC) criterion has been used in this review for identification of research question.

##### Population

Adults (above the age of 18 years) irrespective of their ethnicity, origin, gender and socioeconomic status.

##### Concept

Studies from any domain of healthcare such as maternal-child health, hygiene practices, family planning, healthy sexual practices and others.

##### Context

Type of behavior change intervention, setting of intervention delivery, duration and periodicity of intervention, cadre and other characteristics of provider, mode of intervention delivery, characteristics of the communication module.

The following research questions have been identified for the purpose of this scoping review:

a. In which community-based settings have community health workers delivered behavior change interventions?
b. What are the characteristics of different behavior change communication modules used by the community health workers?
c. Which modes of communication have been used for intervention delivery?

#### 2. Identifying relevant studies-Search strategy

The following freely available databases have been chosen by the research team for retrieval of relevant literature: MEDLINE through PubMed, ERIC, JSTOR and ScienceDirect. To ensure optimum retrieval of relevant studies during the ultimate search, one of the researchers conducted pilot searches using pre-identified keywords such as “community health workers” and “behavior change communication”. The search strategy was iterated and modified by identification of additional keywords from the initially retrieved search, and their inclusion in the search string. The search was customized using appropriate combinations of database specific Boolean (AND, OR) and other operators (+, -) and the final search strategy was identified. A pilot search using the final search strategy was performed in the first week of September, 2020. The search strategy for MEDLINE is as follows:

(“behavior change communication”[all fields] OR “behavior change counseling”[all fields] OR “behavior change intervention”[all fields] OR behavior[MeSH terms] OR counseling[MeSH Terms]) AND ((“health worker”[all fields] AND (community[all fields] OR lay[all fields] OR voluntary[all fields] OR volunteer*[all fields])) OR “Allied Health Personnel”[MeSH]) Filters: Clinical Trial Protocol, Comparative Study, Pragmatic Clinical Trial, Randomized Controlled Trial, English

The above search string retrieved 702 results on September 7^th^, 2020.

Screening of the references and bibliography of the included studies will also be done to identify and include more studies in the review.

#### 3. Study Selection

##### The reviewers have formulated the following eligibility criteria for screening

Both qualitative and quantitative studies will be included for the review purpose. The inclusion of studies will be limited to English language only. Editorials, opinions, commentaries, perspectives, guideline and recommendations, viewpoints, book chapters will not be included in the review for the reason of not being primary data. Studies published since the date of inception of selected databases till September 30^th^, 2020 will be included in the review.

A two-stage selection criteria for study selection will be followed. Articles retrieved after the final search will be entered in a reference management software (Endnote or Mendeley) and duplicate articles will be identified and removed. One of the reviewers will first screen the titles and abstracts of the retrieved studies and exclude the studies which do not meet the eligibility criteria. Full-length articles of the studies to be included in the review will be retrieved. A 20% proportion of retrieved studies will be independently screened by another reviewer for fidelity of eligibility criteria and quality control. Uncertainty about inclusion or exclusion of any article will be resolved at this stage itself, if required, with the involvement of third reviewer.

In the later stage, the first reviewer will screen the retrieved full-length articles and exclude the articles not meeting the eligibility. After the decision on full-length articles to be included in the review, one of the reviewers will screen the references of these articles for identification and screening of additional studies for inclusion. Full-length articles identified from second stage and reference screening will be pooled for data extraction.

The whole process of retrieval of studies from databases, screening and reference screening will be illustrated in the form of a PRISMA flow diagram. (Supplementary file 01)

#### 4. Charting the data

A data extraction template will be designed for the study. The data extraction will capture the information in the domains listed in Table 01 (Supplementary file 01).

One reviewer will extract the data from all the studies and enter into this template. Another reviewer will extract and enter data for 20% of the studies randomly drawn from the included studies. Inter-reviewer reliability will be assessed using Kappa statistics and a will be aimed at a minimum of 0.8.

#### 5. Collating, summarizing data and reporting the results

All the reviewers will assess the data and discuss the findings. The extracted data will be presented in the form of descriptive statistics for various the various categories. A narrative approach will be used to describe the findings across all the categories.

##### Ethics and dissemination

The scoping review methodology consists of collection of data from materials available in public domain, ethics clearance is not required. However, this study is being undertaken as a part of a doctoral thesis. Approval for which has been obtained from the institutional ethics committee (IHECPGRPHD067).

The results of this scoping review will be published in the form of journal articles and presented in conferences. We also anticipate that results of this scoping review will be used by policy-makers and public health researchers in formulating intervention modules for effective behavior change.

## Data Availability

The data will be made available from the authors on demand.

