## Supplementary Files for "Influencers of effective behavior change communication interventions delivered by community health workers in adults: A Scoping Review Protocol"

Supplementary file 01.

Figure 01: Flow Diagram of Database Search and Inclusion of Studies.

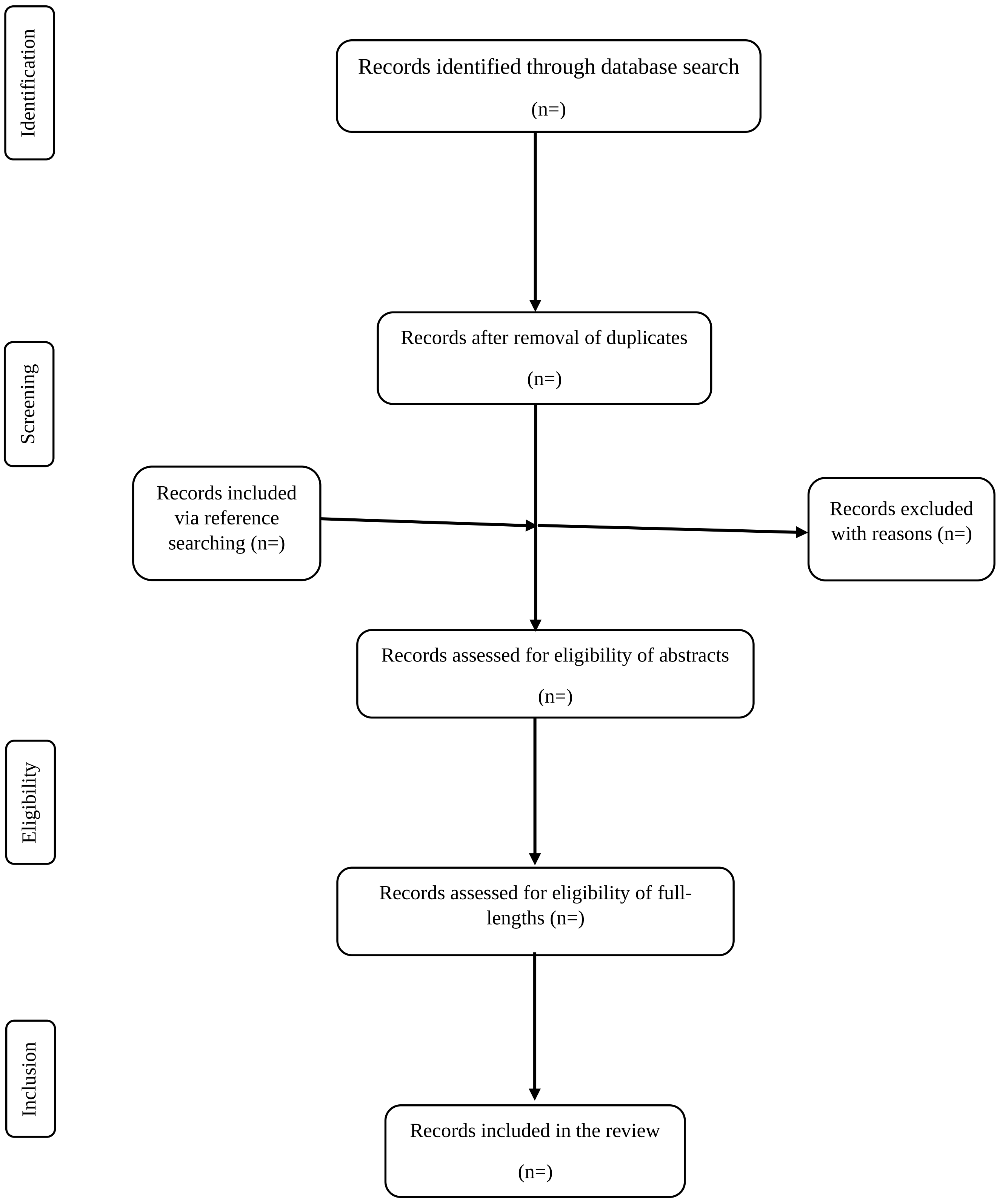

Table 01: Domains/categories for data extraction from selected studies:

| S. No. | Category | Description |
|---|---|---|
| 1 | Bibliography | Title<br>Author<br>Year of publication<br>Journal<br>Place/Country |
| 2 | Community setting | Place of intervention |
| 3 | Intervention type | Personal or grouped |
| 4 | Mode of intervention | Medium used for intervention, human resources, electronic, print and others |
| 5 | Temporal characteristics of intervention | Duration<br>Periodicity |
| 6 | Characteristics of participants | Age group<br>Health condition/behaviour<br>Gender and others |
